# Combining two large clinical cohorts (AIBL and ADNI) to identify multiple lipid metabolic pathways in prevalent and incident Alzheimer’s disease

**DOI:** 10.1101/2020.05.26.20114215

**Authors:** Kevin Huynh, Wei Ling Florence Lim, Corey Giles, Kaushala S Jayawardana, Agus Salim, Natalie A Mellett, Alex Smith, Gavriel Olshansky, Brian G Drew, Pratishtha Chatterjee, Ian Martins, Simon M Laws, Ashley I Bush, Christopher C Rowe, Victor L Villemagne, David Ames, Colin L Masters, Matthias Arnold, Kwangsik Nho, Andrew J Saykin, Rebecca Baillie, Xianlin Han, Rima Kaddurah-Daouk, Ralph N Martins, Peter J Meikle

## Abstract

Changes to lipid metabolism are tightly associated with the onset and pathology of Alzheimer’s disease (AD). Lipids are complex molecules comprising of many isomeric and isobaric species, necessitating detailed analysis to enable interpretation of biological significance. Our expanded targeted lipidomics platform (569 lipid species across 32 lipid (sub)classes) allows for detailed isomeric and isobaric lipid separation. We applied the methodology to examine plasma samples from the Australian Imaging, Biomarkers and Lifestyle flagship study of aging (AIBL, n = 1112) and serum from the Alzheimer’s Disease Neuroimaging Initiative (ADNI, n = 800) studies. Cross sectional analysis using both cohorts identified concordant unique peripheral signatures associated with AD. Specific pathways include; sphingolipids, including G_M3_ gangliosides, where their acyl composition drove the major associations, and lipids previously associated with dysfunctional lipid metabolism in cardiometabolic disease including the phosphatidylethanolamine and triglyceride classes. Infomation derived from improved isomeric seperation highlighted pathway-specific changes with ether lipids including plasmalogens implicating perixosmal dysfunction in disease pathology. Longitudinal analysis revealed similar lipid signitures in both AIBL and ADNI cohorts with future disease onset. We utilised the two independent studies to train and validate multivariate lipid models that significantly improved disease classification and prediction. Together our results provide a holistic view of the lipidome and its relationship with AD using a comprehensive lipidomics approach, providing targets for further mechanistic investigation.

## INTRODUCTION

Alzheimer’s disease (AD) is a neurodegenerative disease characterised by progressive decline in cognitive function, usually presenting with memory loss. In the sporadic form of AD, symptoms usually begin to manifest after the age of 65, and with the ageing global population, the number of people with AD has been estimated to reach 81 million worldwide by 2040 ^1^. The failure of many AD clinical trials over recent years has led to the call for a paradigm shift in AD research. It is now recognised that additional underlying mechanisms are involved in the pathogenesis of AD. We seek to provide a deeper molecular understanding of metabolic pathways implicated in AD to identify key enzymes, transporters and signalling molecules that are most amenable for therapeutic targeting. As there no appropriate sporadic mouse models of AD, human studies are essential for better understanding pathogenesis of AD. In particular, statistically powered studies are needed to detect the associations beneath the natural human biological variation.

Lipids are fundamental to every living system. These diverse and biologically important molecules comprise of thousands of individual species, spanning multiple classes and subclasses. In plasma, the majority of lipids are small amphiphilic molecules (including cholesterol) that make up the circulating lipoprotein particles such as high- & low- density lipoprotein (HDL & LDL). With recent advances to mass spectrometry and high-performance liquid chromatography, it is now feasible to examine in detail the comprehensive plasma lipidome in a human population or clinical study^2^. Quantification and characterisation of these diverse lipid molecules form the foundation of the field known as lipidomics.

Evidence that lipids are involved in AD have been demonstrated via alterations observed in phospholipid ^3-5^, plasmalogens^6^, ceramide ^7^, ganglioside ^8^, and sulfatide ^7, 9^ compositions in the brain. Several recent studies indicate that altered phospholipid metabolism associated with AD pathogenesis is also observed in the blood ^5^, ^10^, ^11^, thus encouraging discovery studies for blood-based lipid markers. Furthermore, a recent large-scale genome-wide association meta-analysis has identified genes involved in lipid metabolism as key risk factors for AD ^12^

The plasma lipidome is complex and consists of many isomeric and isobaric species ^13^, these are species that share similar or identical elemental composition but might be structurally different and offer distinct interprations. Existing lipidomic studies often employ techniques that provide poor resolution of these species ^5, 14-17^ limiting their biological interpretation. We have recently expanded our lipidomic platform to better characterise isomeric lipid species, now measuring 569 lipids from 32 classes and subclasses. Our methodology focuses on lipid and lipid-like compounds utilising extensive chromatographic separation. We have applied this method to two independent studies of ageing and Alzheimer’s disease; The Australian Imaging, Biomarkers and Lifestyle (AIBL) flagship study of ageing and the Alzheimer’s Disease Neuroimaging Initiative 1 (ADNI1) cohorts. Here we show the importance of capturing the comprehensive lipidome and highlight the necessity of obtaining molecular structural detail to identify key lipid pathways to link the plasma lipidome with AD and future onset of AD.

## METHODS

### Participants

The AIBL study recruited 1,112 individuals over the age of 60 years into a longitudinal study ^18^ At baseline, this comprised of 768 cognitively normal, 133 with mild cognitive impairment and 211 with AD. Time points for blood/data collection were every 18 months from baseline. Detailed description of the participants was adapted from Ellis *et al*. ^18^ We utilised all available fasted plasma samples (4,106) from baseline up to the 5^th^ time point.

The ADNI1 study started in 2004 and recruited about 800 individuals at baseline. The initial goal was to recruit 200 particpants with mild AD and 200 controls as well as 400 particpants with MCI. Study data analysed here were obtained from the ADNI database, which is freely available online (http://adni.loni.usc.edu/). We utilised serum samples from the ADNI1 baseline cohort.

### Classification of disease state

Classification of MCI and AD in the AIBL cohort has been described extensively in previous publications (*Ellis et al 2009* ^18^). Clinical criteria used to determine disease status included: Mini Mental State Examination score of less than 28, failure on the Logical Memory test (in accordance with the Alzheimer’s Disease Neuroimaging Initiative criteria), other evidence of possible significant cognitive difficulty on neuropsychological testing, a Clinical Dementia Rating score of 0.5 or greater, a medical history suggestive of the presence of illnesses likely to impair cognitive function, an informant or personal history suggestive of impaired cognitive function, or who were consuming medications or other substances that could affect cognition ^18^ A more in-depth description of the ADNI cohort diagnostic criteria is reported elsewhere ^19^, briefly, AD dementia diagnosis was established using NINDS-ADRDA criteria for probable AD. Classification as MCI followed the Petersen et al criteria previously described ^20^.

### Lipid extraction and liquid chromatography mass spectrometry

Lipids were extracted from 10 μL plasma (AIBL) or serum (ADNI), with the addition of an internal standard mix (Supplementary Table 1), using the single phase butanol/methanol extraction method as described previously ^21^. Analysis of plasma extracts was performed on an Agilent 6490 QQQ mass spectrometer with an Agilent 1290 series HPLC system. Mass spectrometry settings and transitions for each lipid class are shown in Supplementary Table 1. Additional experiments using pooled samples were utilised under varying conditions to get acyl composition data. Detailed description of the method and characterisation of lipid composition and identification of isobaric and isomeric species have been described previously ^22^ The additional experiments to characterise the lipid species include acid hydrolysis, fragmentation in the the presence of lithium ions in positive ionisation mode and fragmentation in negative ionisation mode ^22^.

**Table 1.**
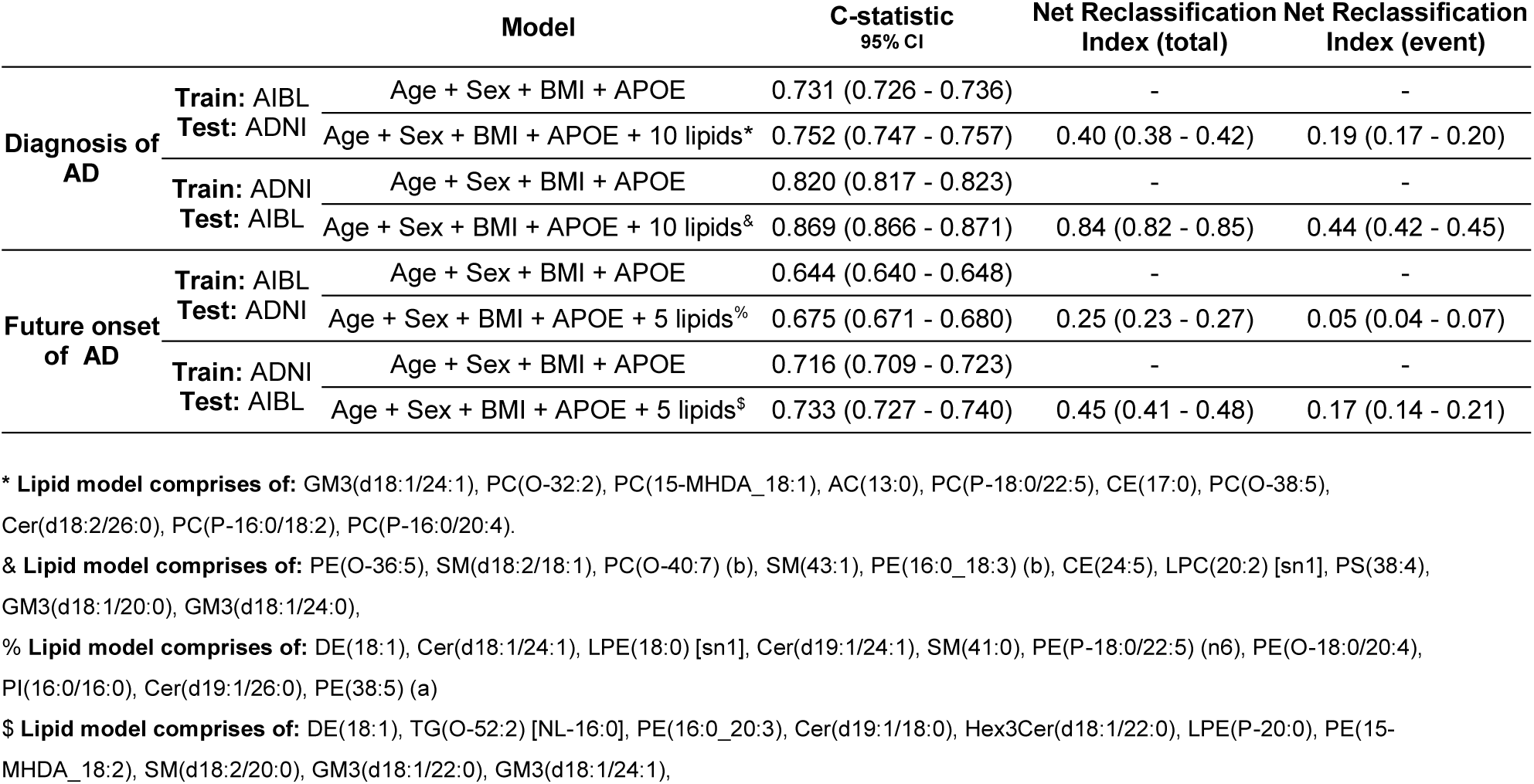
- Summary of modelling statistics.

### Data integration, batch alignment and univariate statistical analysis

Peak area of the lipid species was related to the internal standards to generate concentration data. To remove technical batch variation, the lipid data in each analytical batch (approximately 486 samples a batch) was aligned by using the median value of each lipid of the plasma quality control samples. Final lipid concentrations were log_10_ transformed prior to statistical analysis.

For logistic and Cox regression, lipid data was scaled by the standard deviation and mean centered. Common covariates utilised in statistical analysis include age, sex, BMI, HDL cholesterol, total cholesterol, clinical triglycerides, statin use and omega-3 supplementation. Correction for multiple hypothesis testings was done using Benjamini and Hochberg’s false discovery rate (FDR). Meta-analysis to combine the results between the two cohorts was conducted using an inverse-variance weighted averaged fixed effect meta-analysis.

### Multivariate modelling of Alzheimer’s disease

We leveraged availability of two independent cohorts to train and test several lipid models for currently diagnosed and future incident AD. To define a feature list, in the training cohort we generated a series of models, using forward stepwise regression on top of covariates (to a maximum of 5 lipids per model), minimising Akaike Information Criterion (AIC) under a 10-fold cross validation framework. The frequency of incorporation, after accounting for clustering of co-linear lipids, was used to define features that were used to create a final model in the test set. For testing, the coefficients were generated on 9/10ths of the cohort and tested on the remaining. The concordance statistic (C-statistic) and net reclassification index (NRI) was calculated after 200 repeats. This process was repeated both ways, where AIBL and ADNI were utilised as training and test sets.

### Naming convention of lipid species

The lipid naming convention used here follows the guidelines established by the Lipid Maps Consortium and the shorthand notation established by Liebisch *et al* ^23-25^ We identified several species of interest that are not structurally resolved. These species separated chromatographically but incompletely characterised were labelled with an (a) or (b) to differentiate them, for example PC(P-17:0/20:4) (a) and PC(P-17:0/20:4) (b) where (a) and (b) represent the elution order. Separated isoforms that contain a 16:0 methyl branched fatty acids are presented as MHDA.

## RESULTS

### Lipidomic analysis of the AIBL and ADNI cohort

Between the two cohorts, a total of 5,733 samples (including quality controls and blanks) on 1912 unique individuals were analysed. The characteristics of individuals in the cross-sectional analysis are shown in Supplementary Table 2.

We developed our platform to better characterise the lipids in human plasma ^22^ In total we are able to examine 32 lipid classes and subclasses for both cohorts. In general, a similar correlation structure was observed within the AIBL and ADNI cohorts between lipid classes and many clinical measures (Figure 1). Many commonly reported lipid species associated with AD, such as sphingolipids and ether lipids, are often reported with ambiguous annotations, where the reported species are the sum of several isomers. Here we report the detailed characterisation of these species.

**Figure 1.**
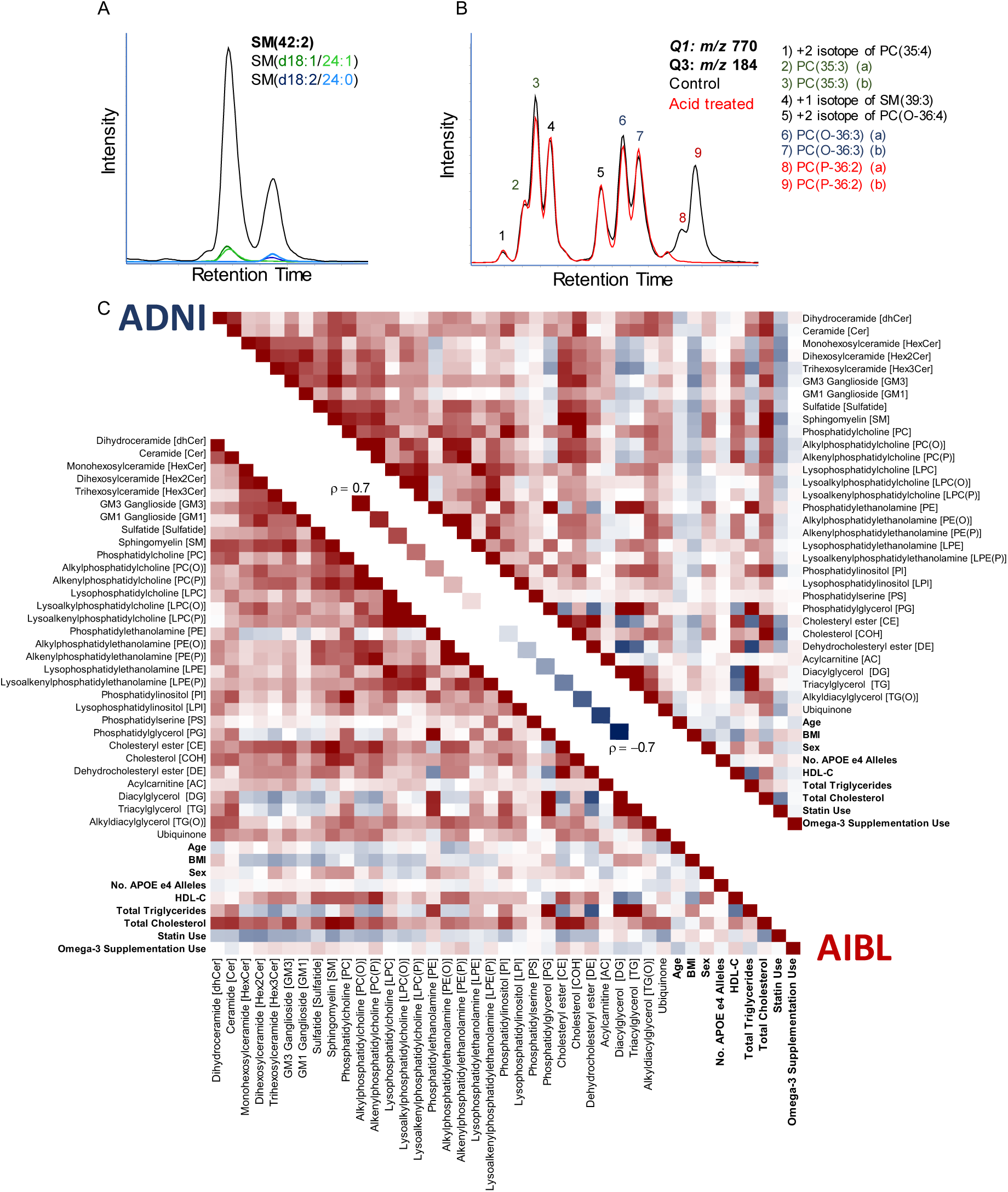
Characterisation of lipid isomeric species and the relationship of lipid classes and subclasses within the AIBL and ADNI cohorts. **A** - Characterisation of sphingomyelin isomers. Black trace corresponds to the chromatogram seen under normal conditions. Additional experimental results in the green and blue traces used for identification, corresponding to SM(d18:1/24:1) and SM(d18:2/24:0) respectively. **B** - Characterisation of glycerophospholipid isomers. Black trace corresponds to the chromatogram seen under normal conditions. Red trace is the same scan after sample acid hydrolysis. **C** - Spearman correlation of total lipid classes, subclasses and commonly reported clinical measures for the AIBL baseline and ADNI studies.

Sphingolipids are structurally resolved through collision induced dissociation (CID), where fragments correspond to the sphingoid base, with the exception of sphingomyelins. Dissociation of sphingomyelin species under normal conditions results in a product ions that yields only sum composition data, i.e. the sphingomyelin species, SM(42:2). To determine sphingomyelin structural composition, we repeated the mass spectrometry analysis on pooled plasma samples in the presence of lithium acetate as described previously ^22^. The lithiated adduct of sphingomyelins produces product ions corresponding to the sphingoid base and n-acyl chain (Figure 1A) allowing for structural identification. Alignment through chromatography highlights that our measurement of SM(42:2) for example, is chromatographically separated into SM(d18:1/24:1) and SM(d18:2/24:0) (Figure 1A).

Similarly, this approach was repeated with the different glycerophospholipid classes to capture isomeric and isobaric structural detail where they were chromatographically resolved. Examination of the transition *m/z* 770.6 / 184.1 corresponding to the phosphatidylcholine / alkylphosphatidylcholine / alkenylphosphatidylcholine species PC(35:3) / PC(O-36:3) / PC(P-36:2) results in 9 distinct peaks (Figure 1B). Here were report complete separation of; diacyl odd-numbered phosphatidylcholine species, the non-plasmalogen ether lipids PC(O) and the plasmalogen ether lipids, PC(P). Results were confirmed by exploiting the susceptibility of plasmalogens to acid hydrolysis (Figure 1B).

### Concordance of associations between two studies with Alzheimer’s disease

After adjustment for covariates (including age, sex, BMI, total cholesterol, HDL-C, triglycerides, site of sample collection, APOE ε4 alleles, omega-3 supplementation, and statin use). There were 12 and 3 classes significantly associated with AD in the AIBL and ADNI1 cohorts respectively after FDR correction, corresponding to 184 and 97 lipids respectively (246 and 197 uncorrected, Figure 3). Meta-analysis using a fixed-effects model identified 218 lipids and 11 classes associated with AD between both cohorts (Figure 2 and 3). The lipid classes associated were predominately from the sphingolipid classes; dihydroceramides (dhCer), trihexosylceramides (Hex3Cer), GM_3_ gangliosides (GM_3_), GM_1_ gangliosides (GM_1_), and ether lipids classes; alkylphosphatidylcholine [PC(O)], alkenylphosphatidylcholine [PC(P)], alkylphosphatidylethanolamine [PE(O)], alkenylphosphatidylethanolamine [PE(P)], alkyldiacylglycerol [TG(O)].

**Figure 2.**
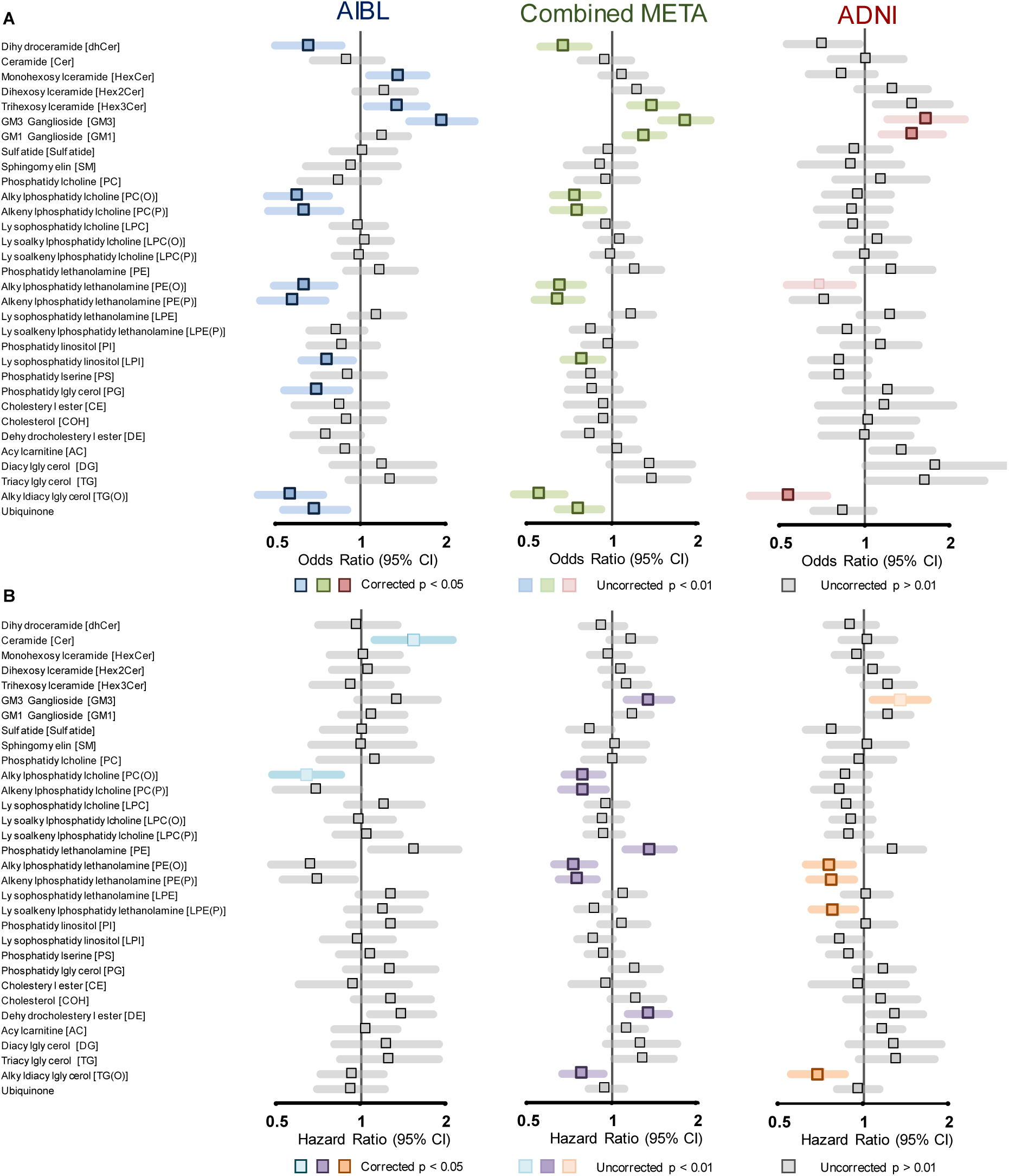
Associations of lipid class totals with prevalent and incident Alzheimer’s disease. Forest plots of lipid class associations for (A) prevalent Alzheimer’s disease and (B) Incident Alzheimer’s disease. Lipid classes are generated by the sum of each individual species measured in each class. Regressions are adjusted (at minimum) for age, sex, BMI, total cholesterol, HDL-C, triglycerides, number of APOE4 alleles, statin use and omega-3 supplementation. Grey squares, not significant, light squares, uncorrected p < 0.01, bolded squares, FDR corrected p < 0.05.

**Figure 3.**
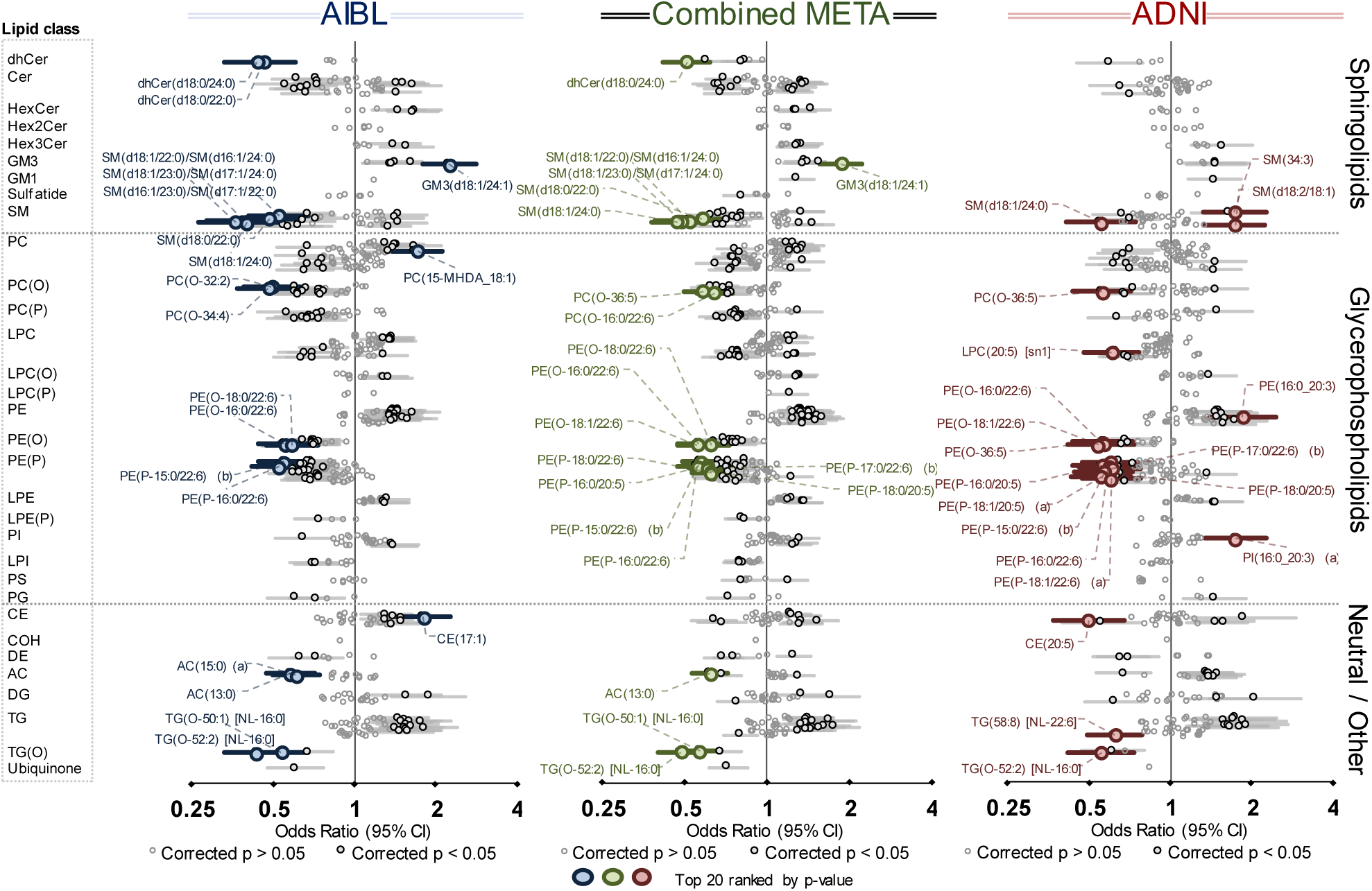
Associations of individual lipid species with prevalent Alzheimer’s disease. Forest plot outlining the logistic regression results of individual species, between controls and prevalent AD in both the AIBL (blue n = 268 cases, 696 controls) and ADNI (red, n = 178 cases, 210 controls) cohorts with the combined meta-analysis in the middle (green). P-value was corrected for multiple comparison using approach by Benjamini and Hochberg. Covariates include age, sex, BMI, total cholesterol, HDLC, triglycerides, number of APOE4 alleles, statin use and omega-3 supplementation. Additional covariates for AIBL include site of blood collection and time point while ADNI includes fasting status. Open circles, not significant, closed dark circles, significant after FDR correction, coloured circles, top 20 associations ranked by p-value.

While all plasmalogens (alkenyl classes) are ether lipids, not all ether lipids are plasmalogens. This distinction is important when factoring in their biological significance. Independent of multiple covariates, the majority of the ether lipid classes were negatively associated with AD (Figure 2 and 3A). This effect is compounded when the ether lipids are esterified with omega-3 fatty acids, such as the 22:6 acyl chain (docosahexaenoic acid, DHA). However, it should be noted that ether lipids with polyunsaturated fatty acids other than omega-3 were still negatively associated with AD (Figure 3B, Supplementary Table 3), highlighting that the effect is not necessarily driven by omega-3 fatty acids. A non-glycerophospholipid subclass of ether lipids, alkyldiacylglycerols, TG(O), were negatively associated with AD, despite the species measured predominately containing saturated or monounsaturated species (Figures 2 and 3).

Ceramide and sphingomyelin species presented with both positive and negative associations with AD resulting in no significant associations at the class level. In both cohorts, dhCer was notability negatively associated with AD. Negative associations were primarily driven by 22:0 and 24:0 saturated species in the sphingolipid n-acyl moiety, while positive associations were predominately the shorter 18:0, 20:0 and monounsaturated 24:1 species. The sphingoid base had no apparent influence on the association in AD. This sphingolipid pattern in AD is much weaker in the ADNI1 cohort (Figure 3, Supplementary table 3).

This effect was also seen for other complex sphingolipids, where a positive association or trend was observed at the class level (monohexosylceramide, HexCer, dihexosyolceramide, Hex2Cer, Hex3Cer, GM_3_ and G_M1_ gangliosides) in both AIBL and ADNI cohorts (Figure 2), but individual species within these classes present with the same opposing relationship (negative association with the n-acyl chains 22:0, 24:0, positive association with the 18:0, 20:0 and 24:1 species). This combined trend resulted in no association with 22:0 and 24:0 n-acyl sphingolipids but an increased association in sphingolipids with n-acyl chains 18:0, 20:0 and 24:1, such as G_M3_(d18:1/24:1) which has the strongest positive association with AD (Figure 3, Supplementary table 3).

A notable pattern observed in both AIBL and ADNI cohorts, even after adjustment for clinical measures of cholesterol and triglycerides, was the positive associations of the lipid classes phosphatidylethanolamine, PE, and triglyceride, TG (Figure 3). This effect has been noted in diseases where dyslipidemia is prevalent such as T2D ^26^.

### Similar lipids are associated with cross sectional and longitudinal analysis of Alzheimer’s disease in both cohorts

We explored using cox regression models lipids associated with the risk of developing AD in the future. Baseline characteristics of this analysis are presented in Supplementary Table 2. After adjustment for covariates (age as time scale, sex, BMI, total cholesterol, HDL-C, triglycerides, site of sample collection, APOE ε4 alleles, omega-3 supplementation and statin use), there were 72 species associated with incident AD in the meta-analysis (Figure 4, Supplementary Table 4). 161 lipids had uncorrected p-values < 0.05 and the majority of these show the same direction of association as observed for AD (Figure 4A). These include individual species from the ether lipids, sphingolipids, phosphatidylethanolamine, and triglyceride classes.

**Figure 4.**
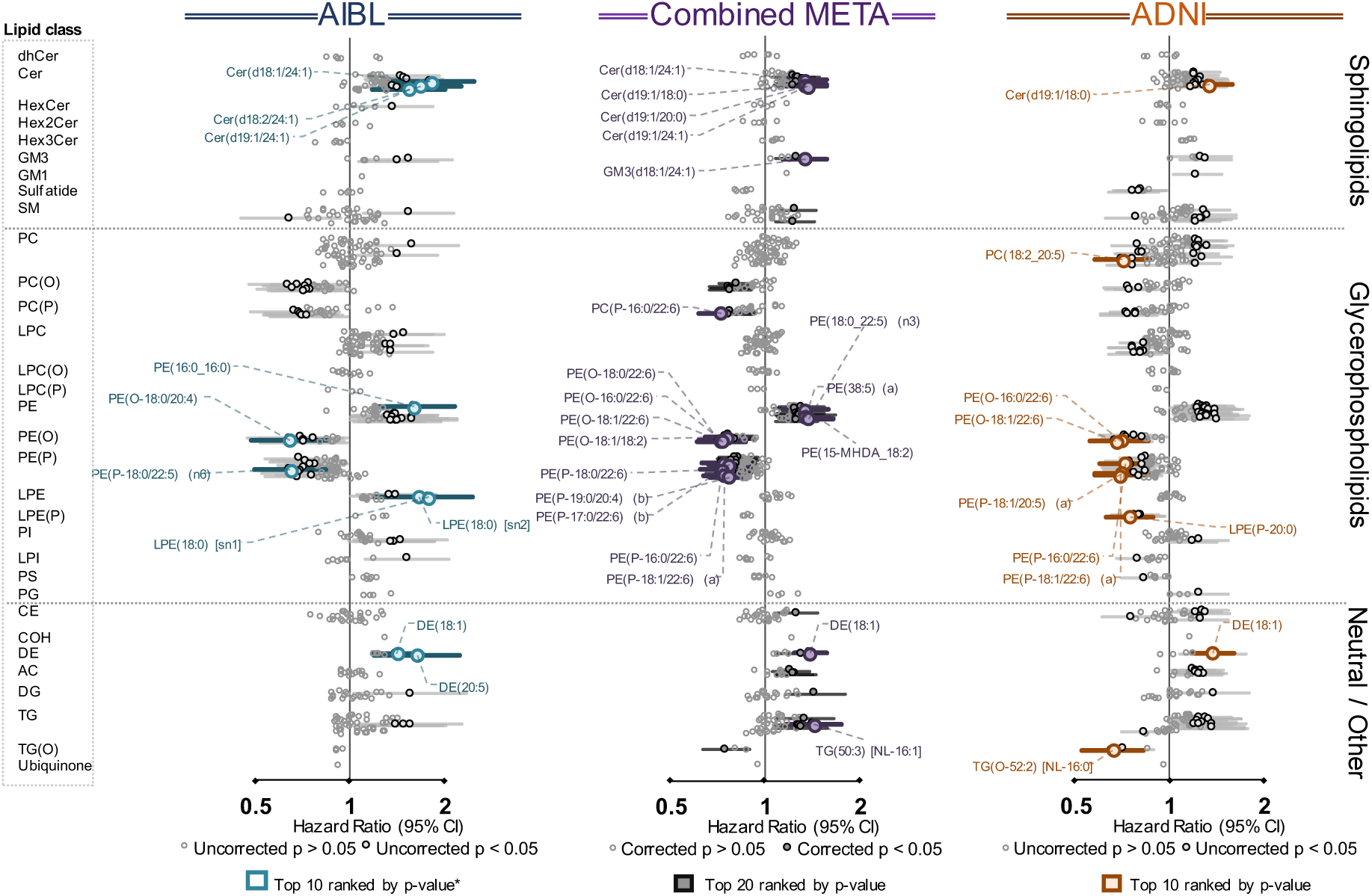
Associations of individual lipid species with future onset Alzheimer’s disease. Forest plot outlining the logistic regression results of individual species, between non-converters and future converters in both the AIBL (light blue n = 68 cases, 714 controls) and ADNI (red, n = 166 cases, 397 controls) cohorts with the combined meta-analysis in the middle (green). P-value was corrected for multiple comparison using approach by Benjamini and Hochberg. Covariates include age (set as timescale), sex, BMI, total cholesterol, HDL-C, triglycerides, number of APOE4 alleles, statin use and omega-3 supplementation. Open circles, not significant, closed dark circles, uncorrected p-value, coloured squares, top 10/20 associations ranked by p-value.

### Multivariate modelling to identify lipids important in diagnosing prevalent or predicting future onset of Alzheimer’s disease

Due to the feature selection process (selecting features based on frequency incorporating into a multivariate model), we observed that informatively prognostic lipids, but are otherwise highly correlated, would frequently be selected once in each model interchangeably, reducing their individual frequency count. We utilised the correlation data to identify lipid clusters of highly correlated species. The frequency of these clusters were then used to rank (sum of individual incorporations), with the most incorporated lipid in each cluster used as the representative for the multivariate models (Supplementary Table 5).

#### Model development in the AIBL, tested in ADNI

In the disease classification model, we observed a final C-statistic of 0.752 (0.747 – 0.757) through the incorporation of 10 lipid species on top of the base model of age, sex, BMI and *APOE* ε4 count (C-statistic of 0.731 [0.726 – 0.736]) with a NRI of 0.40 when tested on the ADNI study. For incident AD, we observed a final C-statistic of 0.675 (0.671 – 0.680), up from 0.644 (0.640 – 0.648) obtained from the base model alone (NRI of 0.40). The summary of results is presented in Table 1.

#### Model development in the ADNI, tested in AIBL

In the parallel analysis, the disease classification model had a final C-statistic of 0.869 (0.866-0.871) through the incorporation of 10 lipid species on top of the base model of age, sex, BMI and *APOE* ε4 count (C-statistic of 0.820 [0.817-0.823]) with a NRI of 0.84 when tested on the AIBL study. For prediction of future disease onset, we observed a final C-statistic of 0.733 (0.727 – 0.740), up from 0.716 (0.709 – 0.723) obtained from the base model alone (NRI of 0.45). The summary of results is presented in Table 1.

## DISCUSSION

Here we present our initial results from the lipidomic analysis of the AIBL and ADNI cohorts. Our goal was to define the complex associations between lipids and AD using our updated methodology by examining each cohort independently and subsequently combining them in a single meta-analysis.The independent nature of these two studies allows for confidence in identifying lipids that are important in AD pathology.

### Multiple lipid classes are positively associated with Alzheimer’s disease

The positive associations of phosphatidylethanolamine, diacylglycerol, and triacylglycerol with AD is similar to the associations seen with both pre-diabetes and type 2 diabetes ^26^. These species tend to correlate closely with clinically elevated levels of triglycerides and reduced HDL cholesterol (dyslipidemia); adjustment for these clinical lipid measures resulted in a loss of significance at the class level; however multiple species within these classes retained a significant association with AD (Figure 3). Interestingly, we observe mostly monounsaturated acyl species driving the associations within these classes. Several studies have highlighted the association of dyslipidemia and insulin resistance with the risk of both vascular dementia and AD ^27, 28^. It may also be possible that the positive association observed with the PE class is a compensatory increase due to the decreases of PE(P) species, a phenomenon that has been reported in plasmalogen-deficient mice ^29^.

### Both alkyl and alkenyl ether lipids are negatively associated with Alzheimer’s disease

A key difference between plasmalogens and non-plasmalogen ether lipids is the vinyl-ether bond in the *sn1* position, which is susceptible to oxidation ^30^. Little of the recently reported metabolomic literature with respect to AD has differentiated these two classes ^5,14-17^. To further complicate this issue, studies which have obtained data from unit resolution mass spectrometers were unable to differentiate these species from isobaric species (odd numbered di-acyl lipids), unless extensive chromatography has also been employed. For example, PC(35:2) has a mass difference of 0.036 Da from PC(O-36:2) and PC(P-36:1), of which the latter two share identical masses (isomeric). When measured on a unit-resolution instrument without chromatography ^5^, these all contribute to the same signal. This can raise issues in instances where these species associate differently with the outcome. This is exemplified in the inverse relationship of odd-chain and ether lipids with AD (Supplementary Table 3), highlighting the importance of differentiating these species.

It has been proposed that plasmalogens act as endogenous antioxidants through their vinyl-ether bond and that increased oxidative stress may explain the lower levels observed in AD patients ^31^. However, both plasmalogen and non-plasmalogen ether species showed similar associations with AD. Alkyl ether lipids are less susceptible to oxidation compared to their plasmenyl counterparts, suggesting associations with AD more likely reflect changes to the biosynthetic pathway. In support of this, Grimm et al. reported dysregulation of the peroxisomal enzymes relating to plasmalogen synthesis in AD ^32^. Plasmalogens have been reported to be involved in several physiological functions including maintenance of lipid raft domains ^33^ which are important for secretase function, responsible for Aβ production ^34^, cholesterol efflux ^35^ and cellular survival ^36^. Importantly, peripheral intravenous administration of plasmalogens has been shown to inhibit Aβ accumulation in a study involving neuroinflammation ^37^. Impairment to the synthetic pathway of ether lipids may have deleterious downstream effects, and, in fact, these species have been proposed as potential therapeutic compounds ^38^.

### Sphingolipids highlight complex acyl associations with Alzheimer’s disease

We have observed diverse associations of specific n-acylated ceramides with AD: positive associations were observed with species containing 18:0, 20:0 and 24:1 fatty acids; whereas negative or neutral associations were observed for species containing 22:0, 24:0 and 26:0 fatty acids, irrespective of the sphingoid base. In contrast, significant associations of sphingolipids with incident AD were only observed for species containing nervonic acid (24:1). This effect was much stronger in the AIBL study, while ADNI exhibited similar trends but were not significant after FDR correction. Despite this, the meta analysis of sphingolipid associations resulted in much more power than utilising the AIBL study alone (Supplementary table 3).

Synthesis of 24:1 is likely through elongation of 18:1 which itself is synthesized through unsaturation via stearoyl-CoA desaturase 1 (SCD-1). Interestingly, increased SCD-1 activity has previously been associated with AD ^39^, and this may be contributing to the increased abundance of these monounsaturated lipid species. We observed similar positive associations with 18:1 relative to 18:0 in many of our measured lipid species, particularly species with a single fatty acid (e.g. LPC and CE species, Supplementary Table 3). Interestingly SCD-1 is associated with insulin resistance and adiposity, and mouse experiments have shown that disrupting SCD-1 function can potentially reduce body adiposity and improve insulin sensitivity ^40^. This is relevant as obesity, insulin resistance and diabetes type II are all likely linked to increased risk of AD ^41^

Specific ceramide synthases are responsible for the n-acylation of ceramide species, in particular, ceramide synthase 2 (20:0 to 26:0) and 3 (22:0 to 26:0) ^42^. Decreases in the activity of ceramide synthase 2 (CerS2) have been observed in AD, early in pathogenesis ^43^. Thus the negative association of Cer(22:0), Cer(24:0) and Cer(26:0) may be driven by decreases of CerS2; while the positive association of Cer(24:1) driven by SCD-1, cancels out the negative association observed with CerS2 which would, on its own, be expected to result in decreased levels of this species.

Gangliosides are a group of sphingolipids with oligosaccharide groups linked to the sphingoid base. GM_3_ gangliosides, the most abundant circulating ganglioside class, is positively associated with AD. Gangliosides have been reported to accelerate Aβ aggregation, leading to deposition in the brain ^44^. While the mechanism leading to increased circulating gangliosides is currently not known, gangliosides are not commonly measured, and thus far no other reports have described an association between circulating GM_3_ gangliosides and AD.

### Multivariate modelling of lipids in Alzheimer’s disease

We were able to identify several lipids that appear to be key in identifying individuals that have, or are at risk of developing AD. These lipids highlight unique features that provide information on top of easily obtained anthropometric (age, sex, BMI) and biochemical features (*APOE* ε4 alleles). While we were able to test and validate these models across the two cohorts, ultimately a population study will be required to fully assess model performance.

### Concluding remarks

We have performed one of the most comprehensive lipidomic analysis of AD to date by utilising two large, independent clinical studies in AD. We have provided a holistic picture of lipid dysregulation associated with both prevalent and incident AD. Our plasma lipid dataset expands the scientific literature by providing greater resolution and allowing fine-granular analysis of the lipidome. Here we have highlighted specific changes to the ether lipid pathway, where plasmalogens are not the only drivers of ether lipid associations. Class-wide and species-specific changes highlight the necessity of a broad and detailed assay to capture these minute differences in the lipidome. We have demonstrated the potential of plasma lipids as the markers to improve assessment of prevalent and incident AD, highlighting the importance of these small molecules in both disease prognostics and understanding the metabolic changes occurring with the disease.

## Data Availability

Data to support this finding is available online and upon request. The entire ADNI lipidomic and clinical characteristic data is available online (adni.loni.usc.edu) and the remaining data (ABIL) used in this study is available from both the corresponding authors and online at https://aibl.csiro.au upon reasonable request.

## ACKNOWLEDGEMENTS

Funding for the AIBL study was provided in part by the study partners [Commonwealth Scientific Industrial and research Organization (CSIRO), Edith Cowan University (ECU), Mental Health Research institute (MHRI), National Ageing Research Institute (NARI), Austin Health, CogState Ltd.]. The AIBL study has also received support from the National Health and Medical Research Council (NHMRC) and the Dementia Collaborative Research Centres program (DCRC2), as well as funding from the Science and Industry Endowment Fund (SIEF) and the Cooperative Research Centre (CRC) for Mental Health – funded through the CRC Program (Grant ID:20100104), an Australian Government Initiative. The authors thank the AIBL study participants for their contribution to the study. K.H was supported by a Dementia Australia Research Foundation Scholarship. PJM is supported by a Senior Research Fellowship from the National Health and Medical Research Council of Australia. Support for the metabolomics sample processing, assays and analytics reported here was provided by grants from the National Institute on Aging (NIA); NIA supported the Alzheimer’s Disease Metabolomics Consortium which is a part of NIA’s national initiatives AMP-AD and M^2^OVE-AD (R01 AG046171, RF1 AG051550, RF1 AG057452, and 3U01 AG024904-09S4). Additional NIH support from the NIA, NLM and NCI for analysis includes P30 AG10133, R01 AG19771, R01 LM012535, R03 AG054936, R01 AG061788, K01 AG049050 and R01 CA129769. M.A. is supported by National Institute on Aging grants RF1 AG057452, RF1 AG058942, RF1 AG059093, and U01 AG061359. M.A. is also supported by funding from Qatar National Research Fund NPRP8-061-3-011. K.N is supported by NLM R01 LM012535 and NIA R03AG054936.This work was also supported in part by the Victorian Government’s Operational Infrastructure Support Program.

Data collection and sharing for the Alzheimer’s Disease Neuroimaging Initiative (ADNI) was supported by National Institutes of Health Grant U01 AG024904. ADNI is funded by the National Institute on Aging, the National Institute of Biomedical Imaging and Bioengineering, and through generous contributions from the following: AbbVie, Alzheimer’s Association; Alzheimer’s Drug Discovery Foundation; Araclon Biotech; BioClinica, Inc.; Biogen; Bristol-Myers Squibb Company; CereSpir, Inc.; Cogstate; Eisai Inc.; Elan Pharmaceuticals, Inc.; Eli Lilly and Company; EuroImmun; F. Hoffmann-La Roche Ltd and its affiliated company Genentech, Inc.; Fujirebio; GE Healthcare; IXICO Ltd.;Janssen Alzheimer Immunotherapy Research & Development, LLC.; Johnson & Johnson Pharmaceutical Research & Development LLC.; Lumosity; Lundbeck; Merck & Co., Inc.;Meso Scale Diagnostics, LLC.; NeuroRx Research; Neurotrack Technologies; Novartis Pharmaceuticals Corporation; Pfizer Inc.; Piramal Imaging; Servier; Takeda Pharmaceutical Company; and Transition Therapeutics. The Canadian Institutes of Health Research is providing funds to support ADNI clinical sites in Canada. Private sector contributions are facilitated by the Foundation for the National Institutes of Health (www.fnih.org). The grantee organization is the Northern California Institute for Research and Education, and the study is coordinated by the Alzheimer’s Therapeutic Research Institute at the University of Southern California. ADNI data are disseminated by the Laboratory for Neuro Imaging at the University of Southern California.

This study was only possible with the help of the AIBL research group. Authors that made direct contribution to this study has been listed as authors in this article. Members of the AIBL group that did not participate in the analysis or writing of this report are listed here; https://aibl.csiro.au/about/aibl-research-team/

Part of the data used in preparation of this article were obtained from the Alzheimer’s Disease Neuroimaging Initiative (ADNI) database (adni.loni.usc.edu). Authors that made direct contribution to this study has been listed as authors in this article. As such, the investigators within the ADNI contributed to the design and implementation of ADNI and/or provided data but did not participate in analysis or writing of this report. A complete listing of ADNI investigators can be found at: http://adni.loni.usc.edu/wp-content/uploads/how_to_apply/ADNI_Acknowledgement_List.pdf

Part of the data used in preparation of this article were generated by the Alzheimer’s Disease Metabolomics Consortium (ADMC). Authors that made direct contribution to this study has been listed as authors in this article. Investigators within the ADMC provided data but did not participate in analysis or writing of this report can be found at: https://sites.duke.edu/adnimetab/team/

## CONFLICTS OF INTEREST

A provisional patent has been filed using the AIBL data. Application number: App Number 2018901220; DEMENTIA RISK ANALYSIS; Baker Heart and Diabetes Institute, Edith Cowan University

ABBREVIATONS
Cer: ceramide;
CE: cholesteryl ester;
COH: free cholesterol;
DE: dehydrocholesterol ester;
DG: diacylglycerol;
dhCer: dihydroceramide;
GM1: G_M1_ ganglioside;
GM3: G_M3_ ganglioside;
HexCer: monohexosylceramide;
Hex2Cer: dihexosylceramide;
Hex3Cer: trihexosylceramide;
LPC: lysophosphatidylcholine;
LPC(O): lysoalkylphosphatidylcholine;
LPC(P): lysoalkenylphosphatidylcholine;
LPE: lysophosphatidylethanolamine;
LPE(P): lysoalkenylphosphatidylethanolamine;
LPI: lysophosphatidylinositol;
PC: phosphatidylcholine;
PC(O): alkylphosphatidylcholine;
PC(P): alkenylphosphatidylcholine (PC plasmalogen);
PE: phosphatidylethanolamine;
PE(O): alkylphosphatidylethanolamine;
PE(P): alkenylphosphatidylethanolamine (PE plasmalogen); PG phosphatidylglycerol;
PI: phosphatidylinositol;
PS: phosphatidylserine;
SM: sphingomyelin;
TG: triglyceride;
TG(O): alkyldiacylglycerol.

